# Impact of Diabetes Mellitus and Viral Hepatitis on the Toxicity Profile of Pre-Extensively Drug-Resistant Tuberculosis Chemotherapy: Focus on Neurotoxicity and Ototoxicity

**DOI:** 10.64898/2025.12.02.25341277

**Authors:** Alexey M. Tikhonov, Olga M. Gordeeva, Sergei A. Pavlov

## Abstract

**Objective:** To analyse the impact of comorbid conditions (diabetes mellitus, viral hepatitis) on the frequency and spectrum of adverse drug reactions (ADRs) during the treatment of pre-extensively drug-resistant (Pre-XDR) pulmonary tuberculosis, with a specific focus on mitochondrial toxicity.

**Materials and Methods:** A retrospective cohort study of 247 HIV-negative patients with Pre-XDR tuberculosis was conducted. Patients received chemotherapy regimens containing linezolid, fluoroquinolones, and injectable agents (consistent with guidelines prior to the 2021 WHO update). Toxicity was graded according to CTCAE standards. The correlation between comorbidities and the incidence of neurotoxic and hepatotoxic ADRs was assessed using multivariate regression analysis.

**Results:** High comorbidity burden was identified as an independent predictor of treatment failure (OR=3.45; p=0.007). The cohort was characterized by a high prevalence of viral hepatitis B/C (up to 12.2%) and diabetes mellitus (up to 10.2%). The most frequent ADRs were eosinophilia (33.6%), ototoxicity (26.3%), and polyneuropathy (up to 15.2% in previously treated patients). Surprisingly, age < 40 years was associated with poor outcomes (OR=4.15; p=0.013), likely driven by lower adherence. The development of severe ADRs significantly increased the risk of treatment failure (OR=4.17; p=0.037).

**Conclusion:** The combination of Pre-XDR tuberculosis with metabolic comorbidities creates a synergistic “mitochondrial hit” effect. This manifests as increased neurotoxicity (attributable to linezolid) and ototoxicity. Even in the era of all-oral regimens, the intersection of diabetes and linezolid-based therapy requires enhanced neuroprotective monitoring.

## Introduction

The management of pre-extensively drug-resistant (Pre-XDR) tuberculosis remains a formidable challenge in global health. The epidemic situation is complicated by the increasing prevalence of resistant strains [1, 2]. Despite the introduction of novel drugs such as bedaquiline and delamanid, global treatment success rates hover around 60%, significantly below the WHO targets [3, 4].

A primary driver of treatment failure is the high incidence of adverse drug reactions (ADRs), which often necessitate dose reduction or treatment interruption [5, 6]. This is particularly relevant for regimens containing linezolid and second-line injectables, which are known for their narrow therapeutic indices and mitochondrial toxicity profiles [7, 8]. Furthermore, the evolution of anti-tuberculosis drugs requires constant re-evaluation of safety profiles in clinical settings [9].

Patients with comorbidities represent a highly vulnerable sub-population. It is well-established that diabetes mellitus (DM) and chronic viral hepatitis not only impair immune response but also alter the pharmacokinetics and metabolic handling of anti-tuberculosis drugs [10, 11]. Recent studies have highlighted the complex management required for concurrent diabetes and tuberculosis [12]. However, the specific pathophysiological interactions between these comorbidities and the toxicity profile of modern Pre-XDR regimens are not fully elucidated. Specifically, the hypothesis of cumulative mitochondrial toxicity—where drug-induced mitochondrial inhibition exacerbates pre-existing metabolic mitochondrial dysfunction—requires clinical validation.

### Objective

To analyse the structure of adverse events in patients with Pre-XDR tuberculosis and to evaluate the role of comorbidities (DM, viral hepatitis) in the development of neurotoxic and hepatotoxic reactions.

## Materials and Methods

### Study Design and Population

This retrospective study included 247 HIV-negative patients with Pre-XDR pulmonary tuberculosis treated at the Central Tuberculosis Research Institute (CTRI). The cohort included patients with complex clinical presentations, often requiring surgical interventions or collapse therapy as part of comprehensive treatment [13, 14].

### Definitions

*Pre-XDR Definition:* Consistent with the national and WHO guidelines active during the data collection period (pre-2021 update), Pre-XDR was defined as TB with resistance to isoniazid and rifampicin, plus resistance to any fluoroquinolone and at least one second-line injectable agent (capreomycin, kanamycin, or amikacin) [15].

### Treatment and Monitoring

Patients were stratified into two groups: Group 1 (n=49, newly diagnosed) and Group 2 (n=198, previously treated). Chemotherapy regimens included fluoroquinolones, linezolid, cycloserine, and injectable agents. ADR monitoring included bi-weekly clinical and biochemical assessment during the intensive phase. Ototoxicity was assessed via patient reporting and audiometry where indicated; polyneuropathy was evaluated based on clinical neurological examination. Toxicity severity was graded according to the Common Terminology Criteria for Adverse Events (CTCAE). Adherence to treatment was assessed using the Morisky-Green scale [16].

### Statistical Analysis

Data were analysed using StatSoft STATISTICA 10. Multivariate logistic regression was performed to identify independent predictors of treatment failure. A p-value of < 0.05 was considered statistically significant.

## Results

### 1. Comorbidity Profile

The study population exhibited a significant burden of comorbidities affecting drug metabolism (Table 1). Viral hepatitis B/C was present in 12.2% of newly diagnosed patients and 7.6% of previously treated patients. Diabetes mellitus was diagnosed in 10.2% and 8.1%, respectively.

**Table 1.**
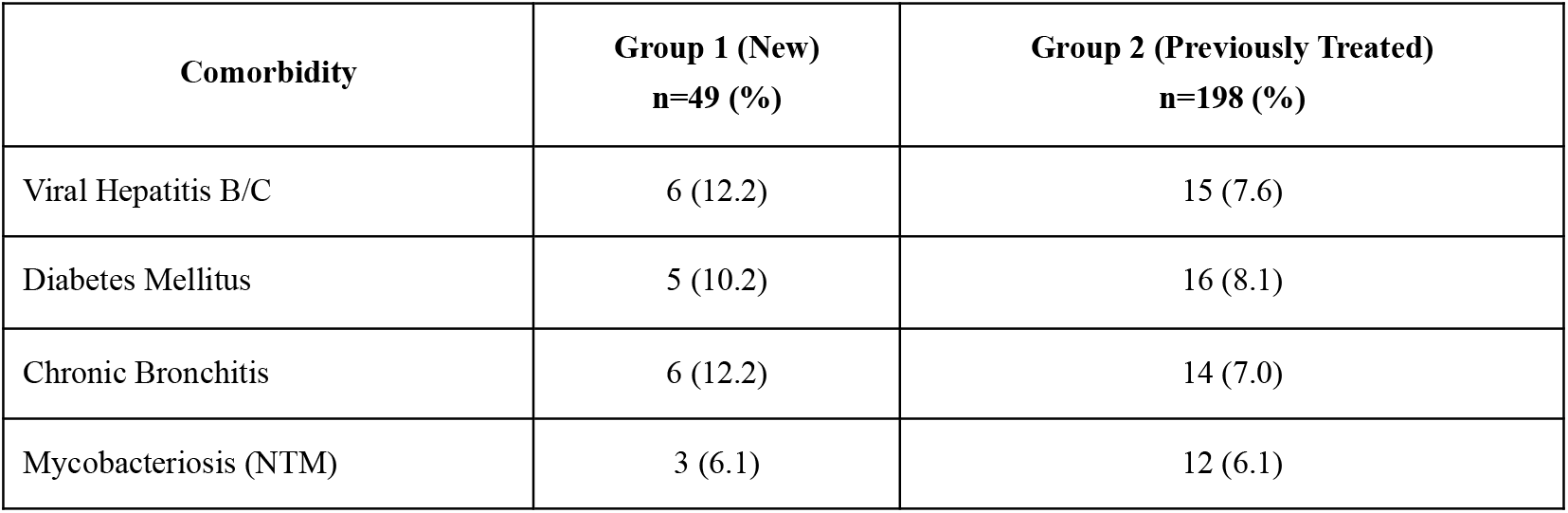
Frequency of key comorbidities in study patients.

### 2. Toxicity Spectrum

The incidence of ADRs was substantial (Table 2). Eosinophilia was the most common laboratory abnormality, affecting approximately one-third of all patients. Clinically significant ototoxicity was observed in 30.6% of Group 1 and 25.3% of Group 2. Polyneuropathy was notably more frequent in previously treated patients (15.2%), likely reflecting cumulative toxicity from prior treatment courses.

**Table 2.** Spectrum of Adverse Drug Reactions (ADRs)

| Adverse Reaction | Group 1 (%) | Group 2 (%) |
| --- | --- | --- |
| Eosinophilia | 32.7 | 33.8 |
| Ototoxicity | 30.6 | 25.3 |
| Nausea / Dyspepsia | 30.6 | 27.8 |
| Polyneuropathy | 8.2 | 15.2 |
| Elevated Transaminases | 22.4 | 20.7 |

### 3. Predictors of Treatment Failure

Multivariate regression analysis (Table 3) identified several independent risk factors. Notably, high comorbidity burden and the development of severe ADRs increased the odds of failure by approximately 3.5 and 4.2 times, respectively.

**Table 3.** Predictors of treatment failure (Multivariate Analysis)

| Factor | Odds Ratio (95% CI) | p-value |
| --- | --- | --- |
| High Comorbidity Burden<br>(Diabetes, Hepatitis, etc.) | 3.45 (1.56 – 5.36) | 0.007 |
| Development of frequent ADRs | 4.17 (1.84 – 7.25) | 0.037 |
| Age < 40 years | 4.15 (2.78 – 6.42) | 0.013 |
| Signs of Lung Destruction | 4.26 (1.86 – 5.72) | 0.012 |

## Discussion

This study highlights the synergistic toxicity between anti-tuberculosis chemotherapy and metabolic comorbidities. Our findings suggest that standard “one-size-fits-all” monitoring protocols may be insufficient for patients with diabetes or viral hepatitis. These results align with recent observations regarding the complexity of treating patients with individually tailored regimens [17].

### The “Mitochondrial Hit” Hypothesis in Diabetic Patients

A key finding is the high rate of polyneuropathy and ototoxicity in our cohort. While the use of injectable agents explains the ototoxicity, the neurotoxicity is likely driven by linezolid. Linezolid inhibits bacterial protein synthesis but also cross-reacts with human mitochondrial ribosomes [18]. Diabetes mellitus independently causes mitochondrial dysfunction and microangiopathy [11, 19]. We postulate a “double hit” mechanism: the drug exacerbates the pre-existing metabolic mitochondrial deficit, lowering the threshold for nerve damage. This is crucial for modern all-oral regimens (such as BPaL/BPaLM), where linezolid remains a backbone drug [20]. In diabetic patients, standard vitamin B supplementation may be inadequate, necessitating earlier dose adjustments or therapeutic drug monitoring (TDM).

### The “Young Patient” Paradox

Our analysis revealed that age under 40 years was a significant predictor of treatment failure (OR 4.15). While older age is typically associated with toxicity, younger age in this specific cohort is likely a proxy for social instability and lower treatment adherence (compliance). Previous studies have identified similar risk factors for multidrug resistance and failure in younger populations [21, 22]. As shown in our compliance analysis (using the Morisky-Green scale), younger patients were more likely to interrupt treatment, which is a known driver of resistance amplification and failure [23].

### Eosinophilia and Hepatotoxicity

The high prevalence of eosinophilia (33%) in a cohort with 7-12% viral hepatitis prevalence is concerning. In this context, persistent eosinophilia should not be dismissed as a minor allergic reaction but investigated as a potential precursor to DRESS syndrome or idiosyncratic hepatotoxicity [24, 25]. In patients with compromised hepatic reserve due to Hepatitis B/C, this serves as an early warning signal.

### Treatment Complexity

Effective management of Pre-XDR TB often requires a multidisciplinary approach, including surgical intervention (resection, collapse therapy). However, our data indicate that the inability to perform surgery is a predictor of failure, consistent with other findings in the field [14, 26].

## Conclusion

1. **Synergistic Toxicity:** Comorbidities (diabetes, hepatitis) are independent predictors of adverse outcomes (OR 3.45), mediated through enhanced drug toxicity.
2. **Relevance to Modern Therapy:** While injectable-induced ototoxicity is decreasing with new WHO guidelines, the linezolid-associated neurotoxicity identified here is of rising importance. Diabetic patients represent a high-risk group for linezolid toxicity due to cumulative mitochondrial stress.
3. **Social Factors:** The association of younger age with treatment failure highlights the critical need for adherence support programs in this demographic.
4. **Clinical Implication:** In comorbid Pre-XDR patients, we recommend intensified monitoring for neuropathy and early investigation of eosinophilia as a marker of systemic hypersensitivity.

## Data Availability

All data produced in the present study are available upon reasonable request to the authors

## References

1. Vasilieva IA, Testov VV, Sterlikov SA. The epidemic situation of tuberculosis in the years of the COVID-19 pandemic - 2020-2021. Tuberculosis and Lung Diseases. 2022;100(3):6–6. [in Russian]

2. Testov VV, Vasilyeva IA, Sterlikov SA, et al. The prevalence of tuberculosis with multiple and broad drug resistance according to the Federal register of persons with tuberculosis. Tuberculosis and Lung Diseases. 2019;97(12):64–64. [in Russian]

3. Global tuberculosis report 2022. Geneva: World Health Organization; 2022.

4. Khawbung JL, Nath D, Chakraborty S. Drug resistant Tuberculosis: A review. Comp Immunol Microbiol Infect Dis. 2021;74:101574.

5. Aslanova AEK, Khasratova KhN. Experience in the use of new anti-tuberculosis drugs in the treatment of tuberculosis patients with widespread drug resistance and evaluation of adverse events. Bulletin of the Central Research Institute of Tuberculosis. 2020;1:99–100. [in Russian]

6. Borisov SE, Dheda K, Enwerem M, et al. Effectiveness and safety of bedaquiline-containing regimens in the treatment of MDR- and XDR-TB: a multicentre study. Eur Respir J. 2017;49(5):1700387.

7. Bahuguna A, Rawat D.S. An overview of new antitubercular drugs, drug candidates, and their targets. Med Res Rev. 2020;40(1):263–263.

8. Stavitskaya NV, Felker IG, Zhukova EM, et al. Multifactorial analysis of the results of the use of bedaquiline in the treatment of MDR/XDR - pulmonary tuberculosis. Tuberculosis and Lung Diseases. 2020;98(7):56–56. [in Russian]

9. Sysoev PG, Liukina AN, Madatova MKK. Evolution of anti-tuberculosis drugs. Avicenna. 2020;64:17–20. [in Russian]

10. Riza AL, Pearson F, Ugarte-Gil C, et al. Clinical management of concurrent diabetes and tuberculosis and the implications for patient services. Lancet Diabetes Endocrinol. 2014;2(9):740–740.

11. Baghaei P, Marjani M, Javanmard P, et al. Diabetes mellitus and tuberculosis facts and controversies. J Diabetes Metab Disord. 2013;12:58.

12. Park SW, Shin JW, Kim JY, et al. The effect of diabetic control status on the clinical features of pulmonary tuberculosis. Eur J Clin Microbiol Infect Dis. 2012;31:1305.

13. Zakharov AV, Tikhonov AM, Polyakova AS, et al. Clinical aspects and effectiveness of complex treatment of pulmonary tuberculosis with a broad drug-resistant pathogen in patients of different registration groups. Bulletin of the Central Research Institute of Tuberculosis. 2022;1:54–68. [in Russian]

14. Medovarov EV, Pavlunin AV, Panchenko NI, et al. Collapse surgery and valvular bronchial blockade in patients with fibrotic-cavernous pulmonary tuberculosis: immediate and long-term results. University Clinic. 2017;4-1(25):119–126. [in Russian]

15. World Health Organization. WHO consolidated guidelines on tuberculosis. Module 4: Treatment - drug-resistant tuberculosis treatment, 2022 update.

16. Morisky DE, Green LW, Levine DM. Concurrent and predictive validity of a self-reported measure of medication adherence. Med Care. 1986;24:67–74.

17. Olaru ID, Lange C, Indra A, et al. High rates of treatment success in pulmonary multidrug-resistant tuberculosis by individually tailored treatment regimens. Ann Am Thorac Soc. 2016;13:1271–1278.

18. Song T, Lee M, Jeon HS, et al. Linezolid treatment for multidrug-resistant tuberculosis. N Engl J Med. 2012;367(16):1508–1508.

19. Garrabou G, Soriano A, López S, et al. Reversible inhibition of mitochondrial protein synthesis during linezolid-related hyperlactatemia. Antimicrob Agents Chemother. 2007;51(3):962–962.

20. Conradie F, Diacon AH, Ngwane N, et al. Treatment of Highly Drug-Resistant Pulmonary Tuberculosis. N Engl J Med. 2020;382(10):893–893.

21. Chen S, Huai P, Wang X, et al. Risk factors for multidrug resistance among previously treated patients with tuberculosis in eastern China: a case-control study. Int J Infect Dis. 2013;17:e1116–20.

22. Kolomiets V, Pavlenko K, Rotenko K. Predictors of the effectiveness of treatment of tuberculosis patients with the development of the COVID-19 pandemic in the region of the Russian Federation. Bulletin of the Academy of Sciences of Moldova. Medicine. 2022;3(74):195–195. [in Russian]

23. Hasker E, et al. Drug-resistant tuberculosis: a survival analysis. Int J Tuberc Lung Dis. 2021;25(3):189–189.

24. Blumenthal KG, Peter JG, Trubiano JA, Phillips EJ. Drug reaction with eosinophilia and systemic symptoms (DRESS) syndrome. Lancet. 2019;393(10175):1016–1016.

25. Tostmann A, Boeree MJ, Aarnoutse RE, et al. Antituberculosis drug-induced hepatotoxicity: concise up-to-date review. J Gastroenterol Hepatol. 2008;23(2):192–192.

26. Gordon AI, Viktorova IB. Problematic issues of surgical treatment of drug-resistant tuberculosis. Bulletin of Modern Clinical Medicine. 2014;7(1):39–39. [in Russian]

